# Validating an Approach for Estimating Appropriate Black Participation in Clinical Trials

**DOI:** 10.1101/2023.07.24.23293100

**Authors:** Jeremy M. Berg, Juan C. Celedón, Naudia N. Jonassaint

## Abstract

Representation of different groups at appropriate levels in clinical trials is of great importance. Factors affecting what appropriate levels include the demographics at the trial sites and the prevalence of the condition under study in different populations. We examined 359 trials published in New England Journal of Medicine, Journal of the American Medical Association (JAMA), and the Lancet in 2020 for information about Black participation rates. Sufficient information for analysis was available in 58 trials. Simulations including both site demographics and prevalence factors revealed that observed Black participation rates were reasonably well correlated with estimated potential Black participation rates, but that actual participation rates were lower than potential rates in 47 out of 58 trials. This approach could be used to estimate appropriate participation rates prior to trial initiation and for analysis of trials upon completion. Promotion of such transparency standards will aid future analyses and should help drive improvements in representation over time.

Clinical trials represent important opportunities to test potential interventions in groups of individuals who can provide meaningful data and who represent populations who might benefit from the trial results. This has been described and highlighted by the recent report “Improving Representation in Clinical Trials and Research: Building Research Equity for Women and Underrepresented Groups” from the United States National Academy of Sciences^1^. One of the overarching conclusions from this report is:

## Improving representation requires transparency and accountability

“Transparency and accountability throughout the entire research enterprise will be critical to driving change and must be present at all points in the research life cycle—from the questions being addressed, to ensuring the populations most affected by the health problems are engaged and considered in the design of the study, to recruitment and retention of study participants, to analysis and reporting of results. Individual investigators and research institutions on the front lines bear responsibility for transparency in reporting progress toward the goals of inclusion in research. Transparency and accountability must also be reinforced by the funding that agencies and industry sponsors have across their portfolios, that regulatory agencies have in their role governing the conduct of research as well as the approval and reimbursement of the drugs and devices that are often the final products of clinical research, and that journal editors and others that disseminate research have in communicating findings.”

The NIH Revitalization Act of 1993 required addressing the inclusion of women and members of minority groups in developing a research design appropriate to the scientific objectives of a given study. This mandate provided an impetus for increasing levels of inclusion of women and minorities in clinical trials but left open for interpretation the term “appropriate”. In almost all cases, concerns have focused on under-representation of minoritized persons trials, but concerns have been raised about over-representation of minorities in phase 1 safety trials^2^ and, recently^3^, about a trial of vitamin D as a potential childhood asthma treatment published in 2020^4^.

An analysis of this term “appropriate” published a decade after the Revitalization Act^5^ discussed three possible goals for inclusion of minorities: (i) “To test hypotheses about possible differences by race or ethnicity”; (ii) “To generate hypotheses about possible differences by race or ethnicity”; and (iii) “To ensure just and equitable distribution of risks and benefits of participation in research”.

To satisfy (i), a trial must have a sufficiently large number of subjects in each racial or ethnic group to be compared, so that statistical power is adequate and the study can yield meaningful results. A trial can satisfy (ii) if it approaches but does not reach the statistical power necessary for (i), allowing exploratory analysis for potential testing in subsequent studies. To satisfy (iii), a trial should include racial and ethnic groups in proportion to the levels to which they are affected by the conditions under study in the populations where the trials are conducted. This goal optimizes the chances that results from the trial will be generalizable to affected populations outside of the trial.

As an example of assessing appropriate participation levels, Loree et al.^6^ conducted an analysis of 230 trials leading to cancer drug approvals from 2008 to 2018. They found that 63% of these trials reported some information about the race or ethnicity of participants and that Blacks were represented at 22% of the rate expected based on cancer incidence in the United States. The analysis did not include racial demographic information specific to the sites where the trials were conducted nor any estimate of the relative prevalence of different cancers by race. This is a substantial limitation since the relative population of Black people often varies substantially by location and most trials largely draw on local communities of subjects.

To extend such analyses to a wide range of conditions and to develop an approach for estimating appropriate levels of different groups in clinical trials, we examined the 359 clinical trials reported in the New England Journal of Medicine, JAMA, and The Lancet in 2020 with a focus on the inclusion of Black trial participants. The emphasis on Black participants is not intended to imply that Blacks are the only group of individuals whose inclusion in clinical trials is important (see, for example,^7^, but rather reflects a pragmatic decision based on the greater frequency of information about Black participation rates in these trials. We identified trials that had information about the number of Black participants and sufficient information about the participating trial sites to allow meaningful inferences about the percentage of Black individuals within the communities served by the sites. The selection of trials is summarized in Figure 1.

**Figure 1.**
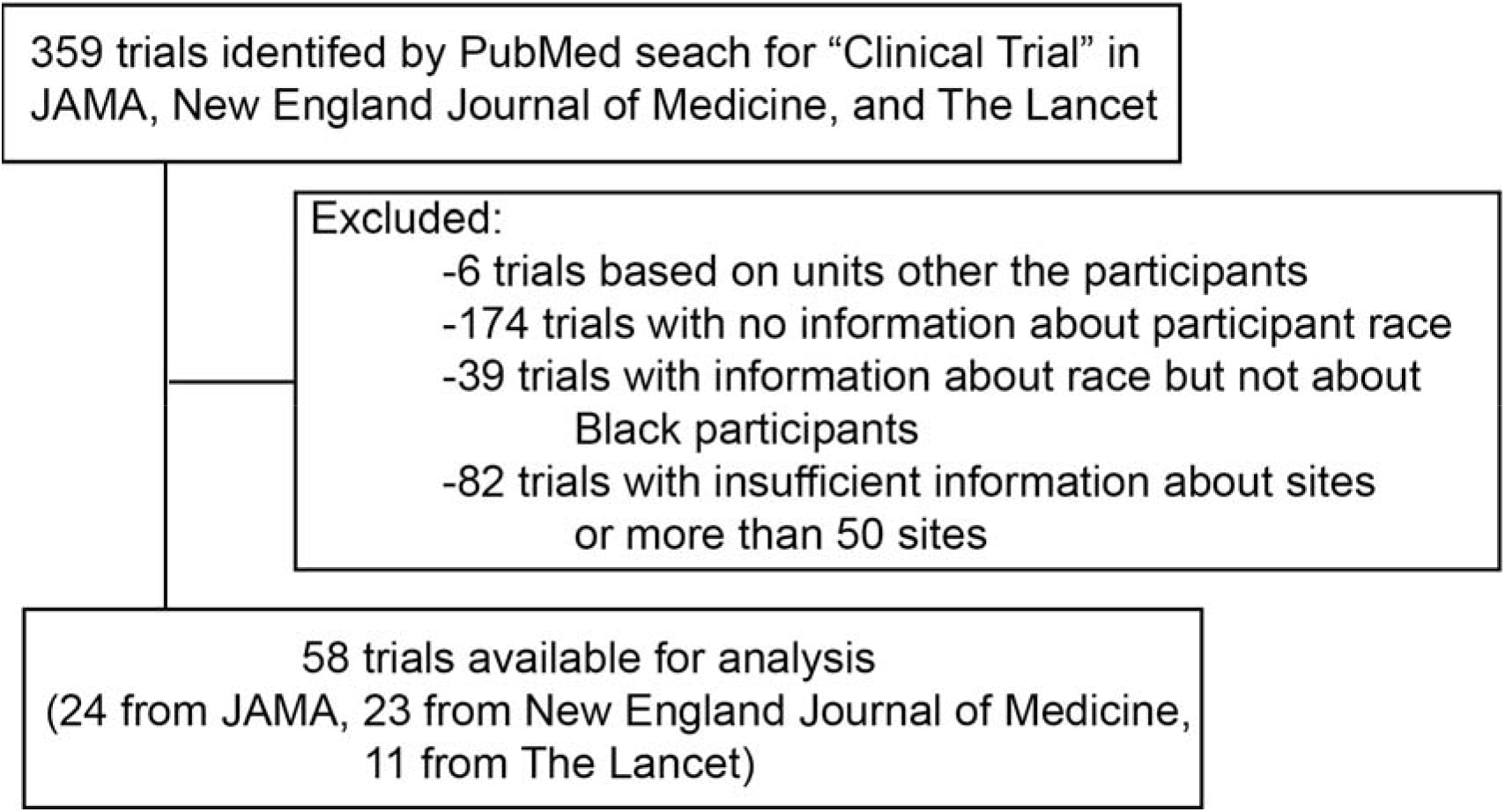
Flowchart for selection of the clinical trials included in this analysis.

Based on these considerations, 58 trials were analyzed. Key parameters for the trials considered are summarized in supplementary information.

## Methods

The percentage of Black participants, if available, was calculated from data in tables, text, or supplementary material from the papers reporting on the selected trials. These percentages can be compared with estimates of the percentage of age-eligible Black individuals who reside at the trial sites and who are affected by the conditions under study. Making such estimates requires (i) identification of the trial sites; (ii) estimates regarding racial demographics at these sites; and (iii) knowledge of or estimates of the fraction of participants recruited at each site.

This approach was implemented as follows. Trials were included if the locations of the sites could be identified from the papers or from associated databases such as ClinicalTrials.gov. Only trials with 50 or fewer sites were included as difficulty in estimation of the availability of potential Black participants increases as the number of sites increases, in the absence of explicit information. Of the 58 trials, 38 took place entirely in the United States, 10 in the United Kingdom, 2 in Canada, 1 in Switzerland, 1 in South Africa, 1 in Australia, and 5 in more than 1 country.

Racial demographics at trial sites were obtained from the United States Census or similar databases for other countries. For sites in the United States, information for both the city and county where the sites were located were included as described below. For each condition, estimates of the relative prevalence for Blacks compared to whites or other populations in the appropriate age group were obtained from literature searches.

For each trial, 1000 simulations were performed. For four trials, information about the number of participants at each site was available from the publication or associated databases and these were used in the simulations. As the number of participants enrolled can vary substantially between sites, these levels were varied from (1/4)(1/n) to 4(1/n) randomly at each site where n is the total number of sites in the trial. The rates of enrollment from cities versus counties at each site in the United States were varied randomly from 25% to 75% since the Black populations in cities and the surrounding counties can vary substantially. Relative prevalence factors were varied randomly based on a standard deviation of 10%. The primary outputs from these simulations were estimates of the percentage of Black participants potentially available for participation across each trial. These results were compared with the observed rates of Black participation with 95% confidence intervals based on simulation variation including estimated sampling errors. All analyses were performed using R [RStudio, Boston, MA, version 1.4.1717 (2021)].

## Results

A comparison of the estimated levels of potential Black participants based on demographics of the trial sites and estimated relative prevalence of the condition(s) under study with the actual Black participation levels are shown in Figure 2.

**Figure 2.**
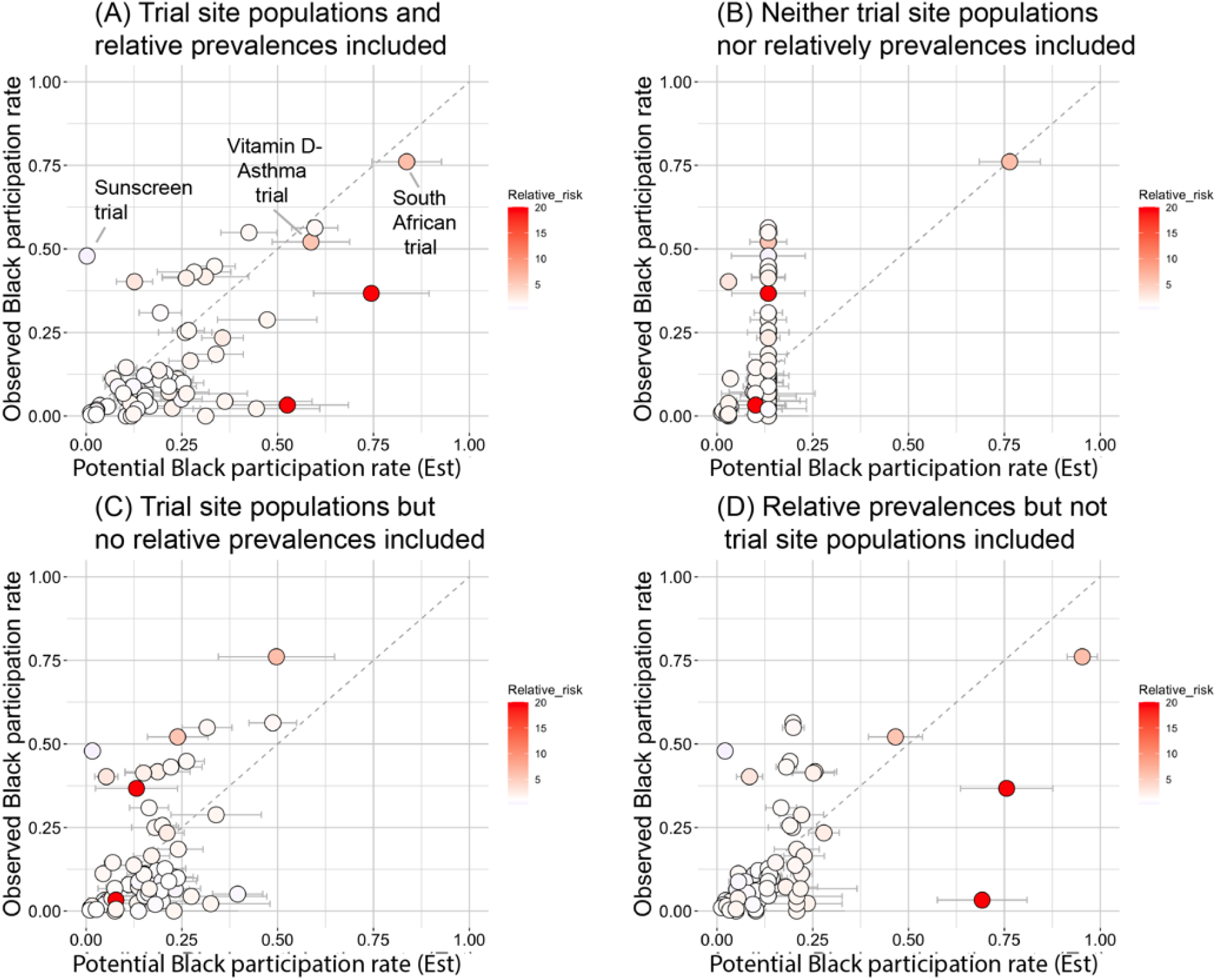
A comparison of the estimated potential Black participation rates and the observed Black participation rate for 58 trials. Estimated 95% confidence intervals are shown. The estimated relative prevalence for Blacks compared to whites for the traits associated with trial participation is depicted by color with deeper red color associated with increased prevalence for Blacks.

One trial related to absorption of sunscreen components intentionally included a range of Fitzpatrick skin types (and hence skin colors) had an estimated potential Black participation rate of < 1% and an observed Black participation rate of 48%^8^. This trial was excluded from subsequent analysis.

These data including the effects of both the Black populations available at trial sites and relative prevalence (Figure 2A) reveal a clear trend with trials with higher estimated available Black populations showing higher levels of Black participation with a Pearson correlation coefficient of 0.70. Both factors (Black population at trial sites and prevalence factor information) are important. Using only the Black population by country and no prevalence factor information (Figure 2B) results in a correlation coefficient of 0.54. This correlation coefficient is reduced to 0.32 if a trial conducted in South Africa is excluded. Adding site-based available Black populations but not prevalence factor information (Figure 2C) increases the correlation coefficient to 0.55. Using the country-based available Black population information and adding the prevalence factor information results in a correlation coefficient of 0.57.

Even with both factors included, the estimated potential Black population is frequently higher than the actual Black participation rate with the actual participation level being less than the estimated available level in 47 out of 58 trials. The estimated potential Black participant level exceeds the observed level by a factor of 2 or more for 23 trials. Thus, this analysis suggests that Blacks appear to be substantially under-represented in most of these trials. Several trials appear to somewhat over-represent Black participants, but further data and analysis are necessary to determine if this is due to uncertainties in the analysis rather than true over-representation. The representation of Blacks in the trial of vitamin D as a potential childhood asthma treatment suggested to overrepresent Black participants appears to be entirely appropriate.

## Discussion

Minority representation in clinical trials is vitally important. Minority recruitment can be compromised by barriers at the individual (e.g., time and resources), interpersonal (e.g., implicit or explicit bias), institutional (e.g., mistrust), and federal (e.g., limited funding]. The conversations over the last 4 decades since the NIH revitalization Act of 1993 has focused on how to remove these barriers and increase the inclusion of women and minority populations into clinical trials^9^. Intentionality and transparency are important components to improving representation. Toward this goal, the New England Journal of Medicine has introduced a requirement for supplementary information describing the representation of study participants in published clinical trials^10^.

Requirements for transparency at the time of publication is an important step. However, explicit requirements for consideration of representation during trial design may also be important for progress. While representation in trials seems to have improved over time, there remains no clear benchmark for what level of representation is “appropriate”. Therefore, next steps should include building models that help researchers understand appropriate representation across racial and other groups prior to study initiation. Calculated benchmarks should consider: 1) the prevalence of disease within racial groups and 2) the makeup of the patient populations from which subjects are being recruited (regional, local and/or clinical). These benchmarks will facilitate improvements in minority representation, study generalizability and impact, and ensure participant safety.

We have described and validated one benchmarking approach based on information about the condition under study and the demographics of the trial sites. This approach would allow estimation of appropriate racial composition of the study population prior to initiation of a clinical trial. The same approach could also be applied after completion of a trial to assess the trial population. Ideally, trial investigators would be transparent about actual participation levels at different sites to facilitate more precise analysis. Funders and publishers could certainly play a role in encouraging performance of such analyses and in data sharing.

As national and international discussions regarding the value of race as a variable continue, we believe that such benchmarks could decrease subjectivity about under-or over-representation of minorities and help the research and clinical communities understand the impact of studies on subpopulations. As a scientific community, study subjects should be assured that their participation and its associated risk is commensurate with the possible benefits to the larger community.

## Data Availability

All data are presented in supplementary material

